# Immune enhancement to prevent infected pancreatic necrosis: A double-blind randomized controlled trial

**DOI:** 10.1101/2021.08.09.21261580

**Authors:** Lu Ke, Jing Zhou, Wenjian Mao, Tao Chen, Yin Zhu, Xinting Pan, Hong Mei, Vikesh Singh, James Buxbaum, Gordon Doig, Chengjian He, Weili Gu, Weihua Lu, Shumin Tu, Haibin Ni, Guoxiu Zhang, Xiangyang Zhao, Junli Sun, Weiwei Chen, Jingchun Song, Min Shao, Jianfeng Tu, Liang Xia, Wenhua He, Qingyun Zhu, Kang Li, Hongyi Yao, Jingyi Wu, Long Fu, Wendi Jiang, He Zhang, Jiajia Lin, Baiqiang Li, Zhihui Tong, John Windsor, Yuxiu Liu, Weiqin Li, Chinese Acute Pancreatitis Clinical Trials Group (CAPCTG)

## Abstract

**BACKGROUND&AIMS:** Infected pancreatic necrosis (IPN) is a highly morbid complication of acute pancreatitis (AP). Since there is evidence of immunosuppression in the early phase of AP, immune enhancement using Thymosin alpha 1 (Tα1), which stimulates both innate and adaptive immunity, may be a therapeutic strategy to prevent IPN. Our aim was to assess the efficacy of early Tα1 treatment on the development of IPN.

**METHODS:** We conducted a multicenter, randomized, double-blind, placebo-controlled trial in patients with predicted severe acute necrotizing pancreatitis (ANP). ANP patients with an APACHE II score≥8 admitted within seven days of the advent of symptoms were considered eligible. Enrolled patients were assigned to receive a subcutaneous injection of Tα1 1.6 mg, every 12 hours for the first 7 days and 1.6 mg once a day for the subsequent 7 days or matching placebo (normal saline). The primary outcome was the development of IPN during the index admission.

**RESULTS:** From Mar 2017 through Dec 2020, 508 patients were randomized at 16 hospitals, of whom 254 were assigned to receive Tα1 and 254 placebo. During the index admission, 40/254 (15.7%) patients in the Tα1 group developed IPN compared with 46/254 patients (18.1%) in the placebo group (difference -2.4% [95%CI -7.4% to 5.0%]; p=0.47). The results were similar in four predefined subgroups. There was no difference in other major complications, including new-onset organ failure (10.6% vs. 15.0%; p=0.15), bleeding (6.3% vs. 3.5%; p=0.15), and gastrointestinal fistula (2.0% vs. 2.4%; p=0.75) during the index admission.

**CONCLUSIONS:** The immune-enhancing Tα1 treatment of patients with predicted severe ANP did not reduce the incidence of IPN during the index admission.

**Trial registration:** Clinicaltrials.gov registry: NCT02473406.

## Introduction

The annual global incidence of acute pancreatitis (AP) is estimated to be 34 per 100,000 individuals ^1^. A smaller subgroup of patients with AP(5-10%) develop acute necrotizing pancreatitis (ANP) ^2^ and can experience a more prolonged disease course and increased morbidity and mortality, especially if infected pancreatic necrosis (IPN) develops ^3, 4^. The bacteria responsible for IPN are often translocated from the gastrointestinal tract and reach the pancreas through several different transmission routes, including hematogenous, lymphatic, and transcoelomic ^5, 6^.

Attempts to reduce the risk of infection in ANP have included the use of prophylactic antibiotics ^7^ and enteral probiotics ^8^. The former is no longer recommended because of issues like antibiotic resistance, methodological quality in previous studies, and fungal superinfection ^9, 10^. The latter is controversial, as a prominent randomized controlled trial found an increased risk of gastrointestinal necrosis associated with probiotic treatment ^8^. Given that there is evidence of immunosuppression in the early phase of AP ^11-14^, a theoretical strategy to reduce the risk of IPN is to boost the host defense (immune enhancement) against bacterial infection ^15^.

Thymosin alpha 1 (Tα1), a polypeptide hormone isolated from the thymus, stimulates both innate and adaptive immunity ^16^. In a pilot study of patients with AP, Tα1was effective in reducing the risk of developing IPN ^17^. Based on this preliminary data, we conducted a multicenter randomized clinical trial to determine the efficacy of early Tα1 treatment on the development of IPN.

## Methods

### Trial design and oversight

This is a multicenter, randomized, double-blind, placebo-controlled, parallel-group trial to assess the efficacy of Tα1 in addition to standard care on the development of IPN in patients with predicted severe ANP. The trial was approved by the local hospital ethics committees of all the participating sites and registered on the ClinicalTrials.gov Registry (NCT02473406) before enrollment commenced. The trial protocol was published in 2020 ^18^. This study was funded by the Science and Technology Project of Jiangsu Province of China (no. SBE2016750187) and partly supported by SciClone Pharmaceuticals Holding Limited, which provided trial drugs and support for meetings during the study period. The funders were not involved in the trial’s design, data collection, interpretation, or manuscript preparation.

### Study population

Patients diagnosed with AP aged 18 to 70 years and with an APACHE II score≥8 and CT severity score ^19^ ≥5 admitted to any of the participating sites within seven days of the onset of abdominal pain were eligible for inclusion. The diagnosis of AP was based on the Revised Atlanta Classification (RAC) criteria ^2^. Patients were excluded if they were pregnant, had a history of chronic pancreatitis, had underlying malignancy, received treatment for pancreatic necrosis prior to enrollment, had a known history of severe cardiovascular, respiratory, renal or hepatic diseases or had pre-existing immune disorders such as AIDS. Detailed inclusion and exclusion criteria are provided in the published protocol ^18^.

At each site, informed consent was obtained from the patients or their next of kin before randomization. Patients were enrolled from Mar 18th, 2017, to Dec 10th, 2020. Follow-up was completed on Mar 10th, 2021.

### Randomization, masking and interventions

Eligible participants were randomized to either the treatment group or the placebo group in a 1:1 ratio. The randomization was stratified by participating sites with a block size of four. Patients were assigned to receive a subcutaneous injection of Tα1 (SciClone Pharmaceutical Co., Ltd, Hongkong) 1.6 mg, every 12 hours for the first 7 days and 1.6 mg once a day for the following 7 days or matching placebo (normal saline, Chengdu Tongde Pharmaceutical Co., Ltd, Chengdu) during the same period. The trial drug was administered for a maximum of 14 days, or until hospital discharge or death, whichever occurred first.

Participants, treating physicians, and investigators were blinded to the treatment allocation to minimize potential bias. The trial statistician was also blinded when developing the statistical programs. Tα1 and placebo were supplied in identically labeled individual vials. All other aspects of the patients’ care were provided on the basis of the international guidelines ^20^. The details for the management of AP are in the published protocol ^18^.

### Data collection

A web-based database (Unimed Scientific Inc., Wuxi, China) was developed for data collection (accessed at capctg.medbit.cn). Before enrollment, a start-up meeting for data entry and storage training was organized at each participating site to ensure high-quality data collection.

### Trial outcomes

The primary outcome was the development of IPN during the index admission. The diagnosis of IPN was made when one or more of the following criteria were present: gas bubbles within pancreatic and/or peripancreatic necrosis on CT; a positive culture from pancreatic and/or peripancreatic necrosis obtained by fine-needle aspiration, drainage, or necrosectomy ^2^. Secondary clinical outcomes include IPN at 90 days after randomization and new-onset organ failure as defined by the Revised Atlanta Classification ^2^ as well as mortality, bleeding requiring intervention, gastrointestinal fistula requiring intervention, positive blood culture, and pancreatic fistula during the index admission. Secondary laboratory outcomes include C-reactive protein (CRP), monocyte human leukocyte antigen-DR (mHLA-DR), and lymphocyte count at day 7 and day 14 after randomization and positive blood cultures. The details and definitions of all outcomes are provided in the published protocol ^18^.

### Statistical analysis

The incidence of IPN during the index admission in our study population was approximately 25% from our previous studies ^21, 22^. A sample size of 520 patients was conservatively estimated to provide 80% power at a 2-sided alpha of 5% to demonstrate an absolute risk reduction of 10% in IPN during the index admission (25% in the placebo group vs. 15% in the Tα1 group) after adding 4% more patients to account for possible dropouts like withdrawal of consent (PASS V.11, NCSS software, Kaysville, USA) ^17^.

Primary analyses were based on the intention-to-treat (ITT) population, and secondary sensitive analyses were done on the per-protocol (PP) population for the primary outcome and key secondary outcomes. Continuous data are reported as means and standard deviations or as medians and interquartile ranges as appropriate, depending on their normality. Categorical data are expressed as numbers and percentages.

The generalized linear model (GLM) was employed to compare group differences in the primary outcome with site as a covariate, and the risk difference, together with its 95% confidence interval, were calculated. Adjusted analyses with prespecified covariates were also performed. The GLM was also employed for analyses of secondary outcomes with treatment as the single predictor. Kaplan-Meier curves were used to compare the cumulative incidence of IPN to 90 days after randomization tested by log-rank test. Four subgroups were predefined for the evaluation of the incidence of IPN during the index admission and 90 days after randomization: the severity of AP (severe and non-severe ^2^), age (>60 and <60 years old), aetiologies of AP (biliary and non-biliary) and extent of pancreatic necrosis (>50% and ≤50%).

Analyses were conducted using SAS 9.4^®^. Statistical tests will be two-sided, and p values <0.05 will be deemed as significant. All authors had access to the study data and had reviewed and approved the final manuscript.

## Results

### Results of recruitment and baseline characteristics

During the study period, 3,569 AP patients were assessed for eligibility, of whom 508 were enrolled in the trial at 16 hospitals in China. Among those 508 randomized patients, 254 were assigned to receive Tα1 and 254 placebo. The most common reasons for exclusion were admission >7 days before evaluation and APACHE score <8. Eleven patients in the Tα1 group and eight patients in the placebo group withdraw consent during treatment but did not refuse follow-up and data usage (**Figure 1**). Three patients in the placebo group stopped research intervention midway due to adverse reactions.

**Figure 1:**
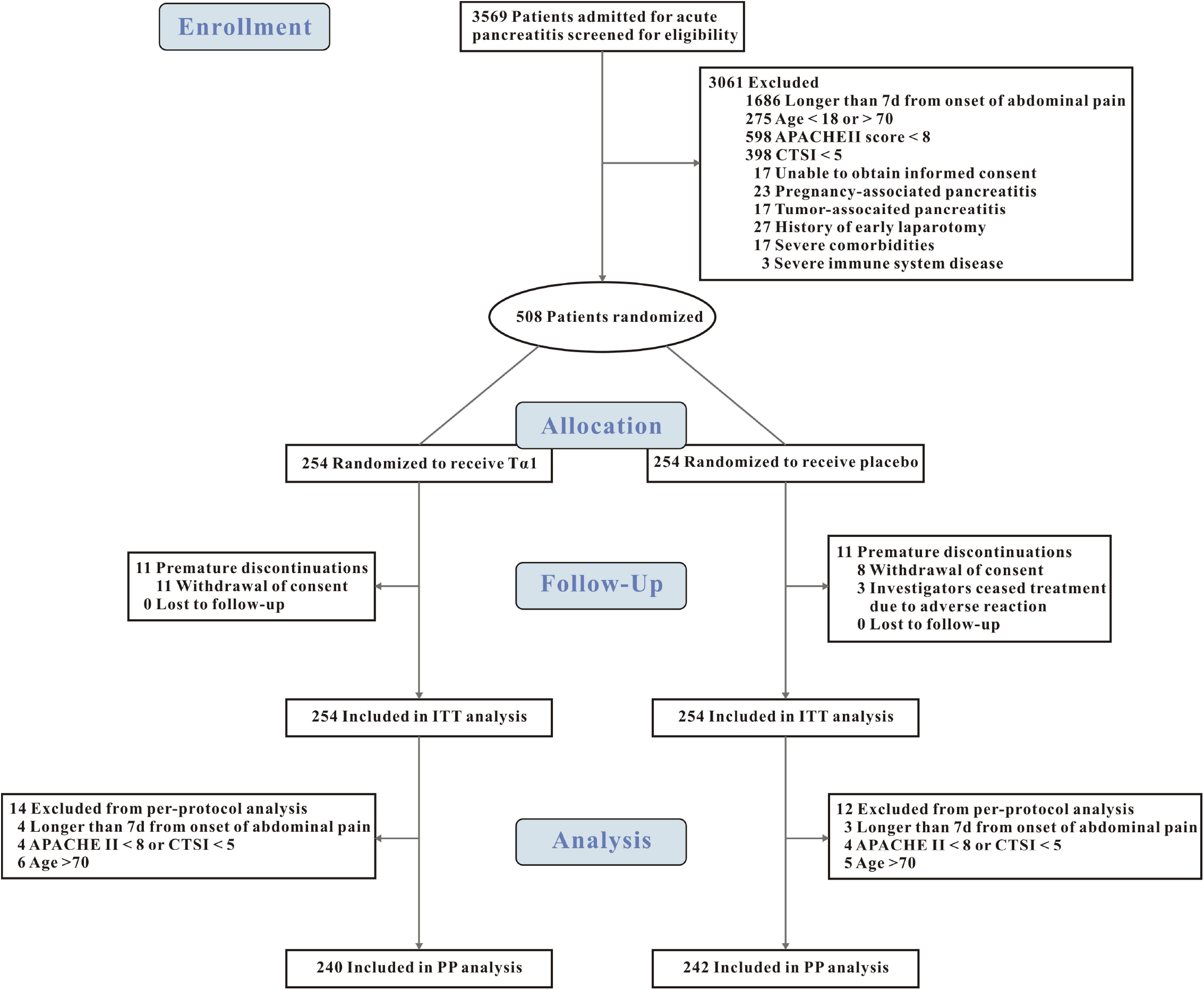
Enrollment, randomization and follow-up of patients in the TRACE trial. TRACE denotes Thymosin α1 in Prevention of Infected Pancreatic Necrosis Following Acute Necrotizing Pancreatitis. APACHE II denotes acute physiology and chronic health evaluation II. CTSI denotes compute tomography severity index. ITT denotes intention to treat. Tα1 denotes Thymosin α1.

Baseline demographics and characteristics were not significantly different between the Tα1 and placebo groups (**Table 1**). In both groups, hypertriglyceridemia was the leading cause of AP, accounting for approximately half of the cases (48.8% vs. 50%).

**Table 1.**
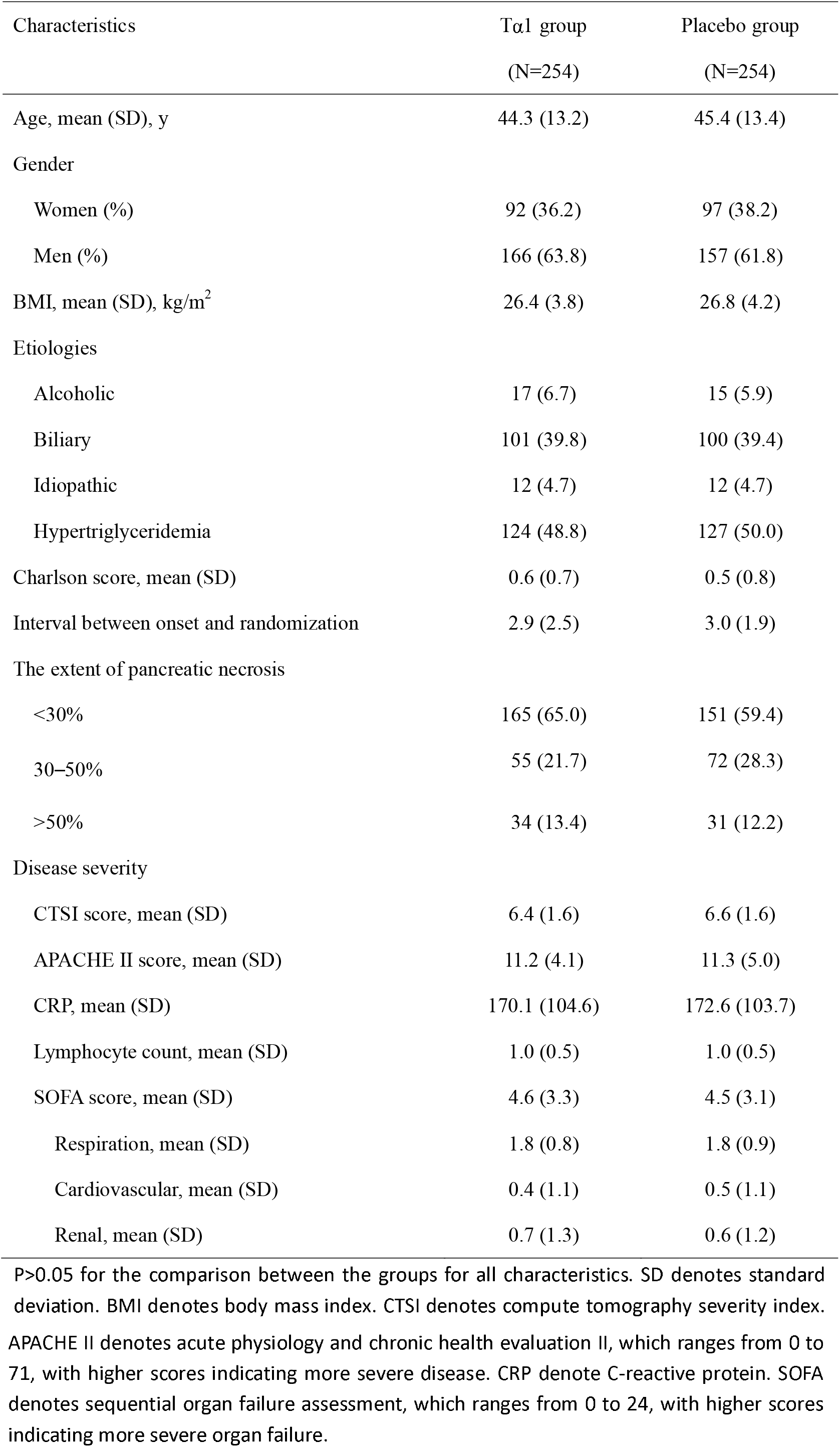
Baseline characteristics of the study subjects

| Characteristics | Tα1 group<br>(N=254) | Placebo group<br>(N=254) |
| --- | --- | --- |
| Age, mean (SD), y | 44.3 (13.2) | 45.4 (13.4) |
| Gender |  |  |
| Women (%) | 92 (36.2) | 97 (38.2) |
| Men (%) | 166 (63.8) | 157 (61.8) |
| BMI, mean (SD), kg/m<sup>2</sup> | 26.4 (3.8) | 26.8 (4.2) |
| Etiologies |  |  |
| Alcoholic | 17 (6.7) | 15 (5.9) |
| Biliary | 101 (39.8) | 100 (39.4) |
| Idiopathic | 12 (4.7) | 12 (4.7) |
| Hypertriglyceridemia | 124 (48.8) | 127 (50.0) |
| Charlson score, mean (SD) | 0.6 (0.7) | 0.5 (0.8) |
| Interval between onset and randomization | 2.9 (2.5) | 3.0 (1.9) |
| The extent of pancreatic necrosis |  |  |
| <30% | 165 (65.0) | 151 (59.4) |
| 30–50% | 55 (21.7) | 72 (28.3) |
| >50% | 34 (13.4) | 31 (12.2) |
| Disease severity |  |  |
| CTSI score, mean (SD) | 6.4 (1.6) | 6.6 (1.6) |
| APACHE II score, mean (SD) | 11.2 (4.1) | 11.3 (5.0) |
| CRP, mean (SD) | 170.1 (104.6) | 172.6 (103.7) |
| Lymphocyte count, mean (SD) | 1.0 (0.5) | 1.0 (0.5) |
| SOFA score, mean (SD) | 4.6 (3.3) | 4.5 (3.1) |
| Respiration, mean (SD) | 1.8 (0.8) | 1.8 (0.9) |
| Cardiovascular, mean (SD) | 0.4 (1.1) | 0.5 (1.1) |
| Renal, mean (SD) | 0.7 (1.3) | 0.6 (1.2) |

P>0.05 for the comparison between the groups for all characteristics. SD denotes standard deviation. BMI denotes body mass index. CTSI denotes compute tomography severity index.
APACHE II denotes acute physiology and chronic health evaluation II, which ranges from 0 to 71, with higher scores indicating more severe disease. CRP denote C-reactive protein. SOFA denotes sequential organ failure assessment, which ranges from 0 to 24, with higher scores indicating more severe organ failure.

### Primary outcome and secondary outcomes

During the index admission, 40/254 (15.7%) patients in the Tα1 group developed IPN compared with 46/254 patients (18.1%) patients in the placebo group (difference -2.4% [95%CI -7.4% to 5.0%]; p=0.47). At 90 days after randomization, 57/254 (22.4%) patients in the Tα1 group developed IPN compared with 65/254 patients (25.6%) in the placebo group (difference -3.3% [95%CI -9.2% to 4.8%]; p=0.39). There was no difference in mortality between groups either within the index admission or at 90 days after randomization (**Table 2**). The Kaplan-Meier curves for the cumulative incidence of IPN until 90 days after randomization are shown in **Figure 2**. There was no significant difference in the probability of developing IPN between the Tα1 and placebo groups (Log-Rank P=0.39).

**Table 2.** Primary and secondary endpoints

|  | **Ta1 group** | **Placebo group** | **Risk difference** | **P value** |
| --- | --- | --- | --- | --- |
|  | **(N=254)** | **(N=254)** | **(95% CI)** |  |
| **Primary endpoint** |  |  |  |  |
| IPN during the index admission, (n, %)** | 40 (15.7) | 46 (18.1) | -2.40 (-7.42, 4.98) | 0.47 |
| **Secondary endpoints\*** |  |  |  |  |
| New-onset organ failure | 27 (10.6) | 38 (15.0) | -4.26 (-8.18, 1.93) | 0.15 |
| Respiratory, (n, %) | 9 (3.5) | 17 (6.7) | -3.15 (-5.08, 1.08) | 0.11 |
| Renal, (n, %) | 10 (3.9) | 7 (2.8) | 1.25 (-1.19, 7.48) | 0.43 |
| Cardiovascular, (n, %) | 14 (5.5) | 20 (7.9) | -2.33 (-5.00, 2.82) | 0.30 |
| Mortality (n, %) | 18 (7.1) | 22 (8.7) | -1.55 (-4.73, 4.21) | 0.52 |
| 90-day mortality (n, %) | 23 (9.1) | 23 (9.1) | 0.03 (-3.79, 6.63) | 0.99 |
| IPN within 90 days after randomization (n, %) | 57 (22.4) | 65 (25.6) | -3.25 (-9.18, 4.83) | 0.39 |
| Bleeding, (n, %) | 16 (6.3) | 9 (3.5) | 2.80 (-0.69, 10.53) | 0.15 |
| Positive blood culture (n, %) | 18 (7.1) | 25 (9.8) | -2.73 (-5.85, 2.83) | 0.27 |
| Gastrointestinal fistula, (n, %) | 5 (2.0) | 6 (2.4) | -0.41 (-1.76, 3.91) | 0.75 |
| Length of ICU stay, mean (SD), d | 14.4 (16.2) | 13.6 (16.4) | 0.75 (-2.07, 3.57) | 0.60 |
| Length of hospital stay, mean (SD), d | 21.0 (21.3) | 20.5 (20.0) | 0.46 (-3.12, 4.03) | 0.80 |
| In-hospital cost, kyuan | 143 (177) | 138 (206) | 5 (-28, 38) | 0.77 |
| **Requirement of invasive intervention** |  |  |  |  |
| Catheter drainage (n, %) # | 36 (14.2) | 39 (15.4) | -1.27 (-6.08, 6.02) | 0.69 |
| Number of drainage procedures, mean (SD) | 0.4 (1.1) | 0.4 (1.2) | -0.02 (-0.21, 0.18) | 0.84 |
| Minimally-invasive debridement (n, %) | 17 (6.7) | 12 (4.7) | 1.96 (-1.44, 8.91) | 0.34 |
| Number of minimally invasive procedures, mean (SD) | 0.2 (1.0) | 0.2 (0.9) | 0.04 (-0.13, 0.21) | 0.61 |
| Open surgery (n, %) | 8 (3.1) | 5 (2.0) | 1.14 (-0.93, 7.37) | 0.41 |

CI denotes confidential interval. IPN denotes infected pancreatic necrosis. ICU denotes intensive care unit. \*All secondary endpoints were registered during the index admission unless otherwise specified; # Both percutaneous or transluminal drainage included; \*\*, adjusted for sites.

**Figure 2:**
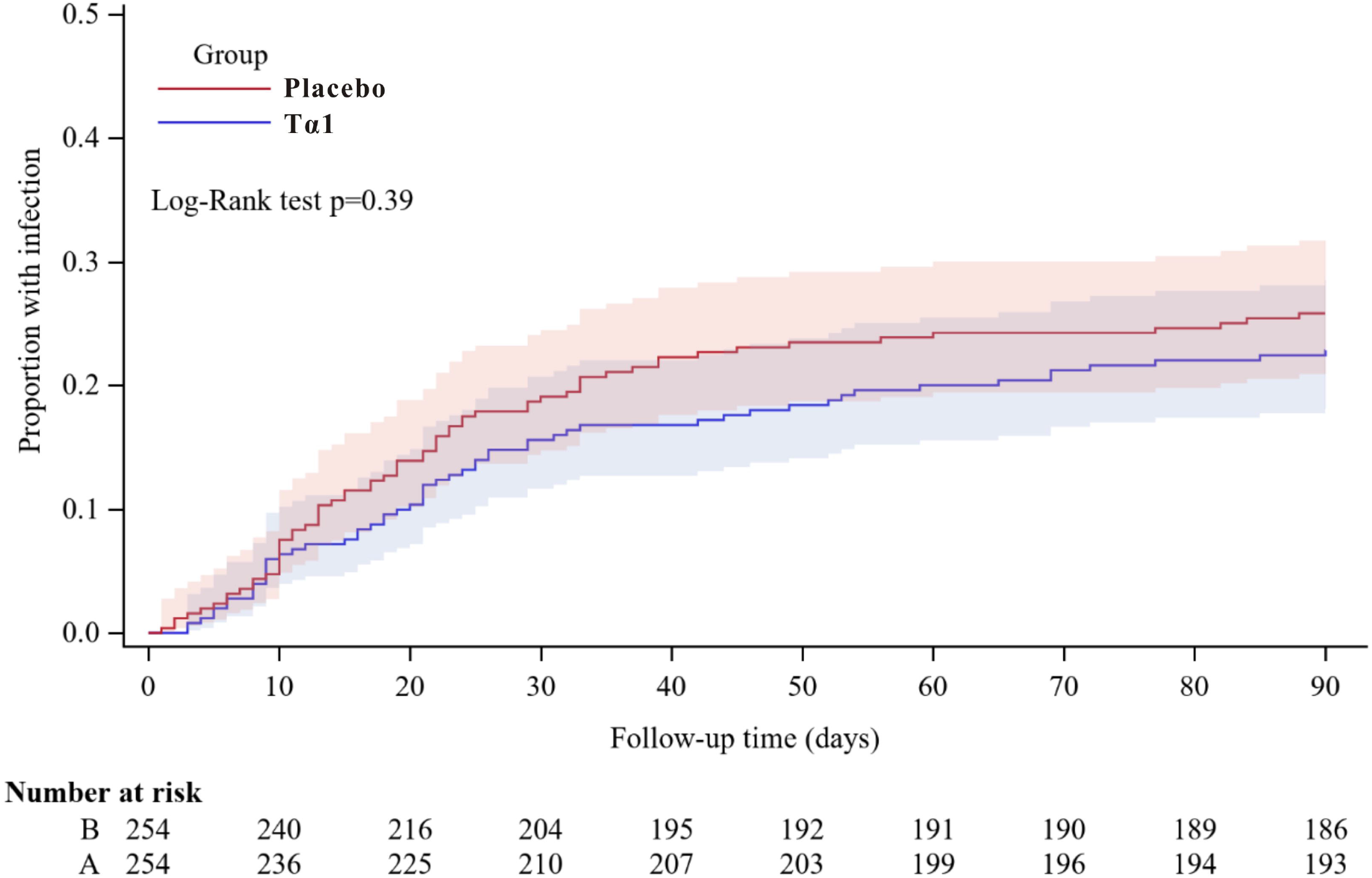
Time-to-infection by day 90 The Kaplan-Meier curves for cumulative incidence of infected pancreatic necrosis from randomization to day 90 in the intention-to-treat population.

There was no difference in other major complications, including new-onset organ failure (10.6% vs. 15.0%; difference -4.3% [95%CI -8.2% to 1.9%]; p=0.15), bleeding (6.3% vs. 3.5%; difference 2.8 [95%CI -0.7 to 10.5]; p=0.15), and gastrointestinal fistula (2.0% vs. 2.4%; difference -0.4% [95%CI -1.8% to 3.9%]; p=0.75) during the index admission. Moreover, there were no significant differences in length of ICU or hospital stay, requirement for catheter drainage and minimally-invasive debridement or open surgery between the patients in the Tα1 and placebo groups (**Table 2**).

### Subgroup analyses

There was no significant heterogeneity in the effect of Tα1 on the incidence of IPN during the index admission and 90 days after randomization in any of the four predefined subgroups (**Figure 3**).

**Figure 3:**
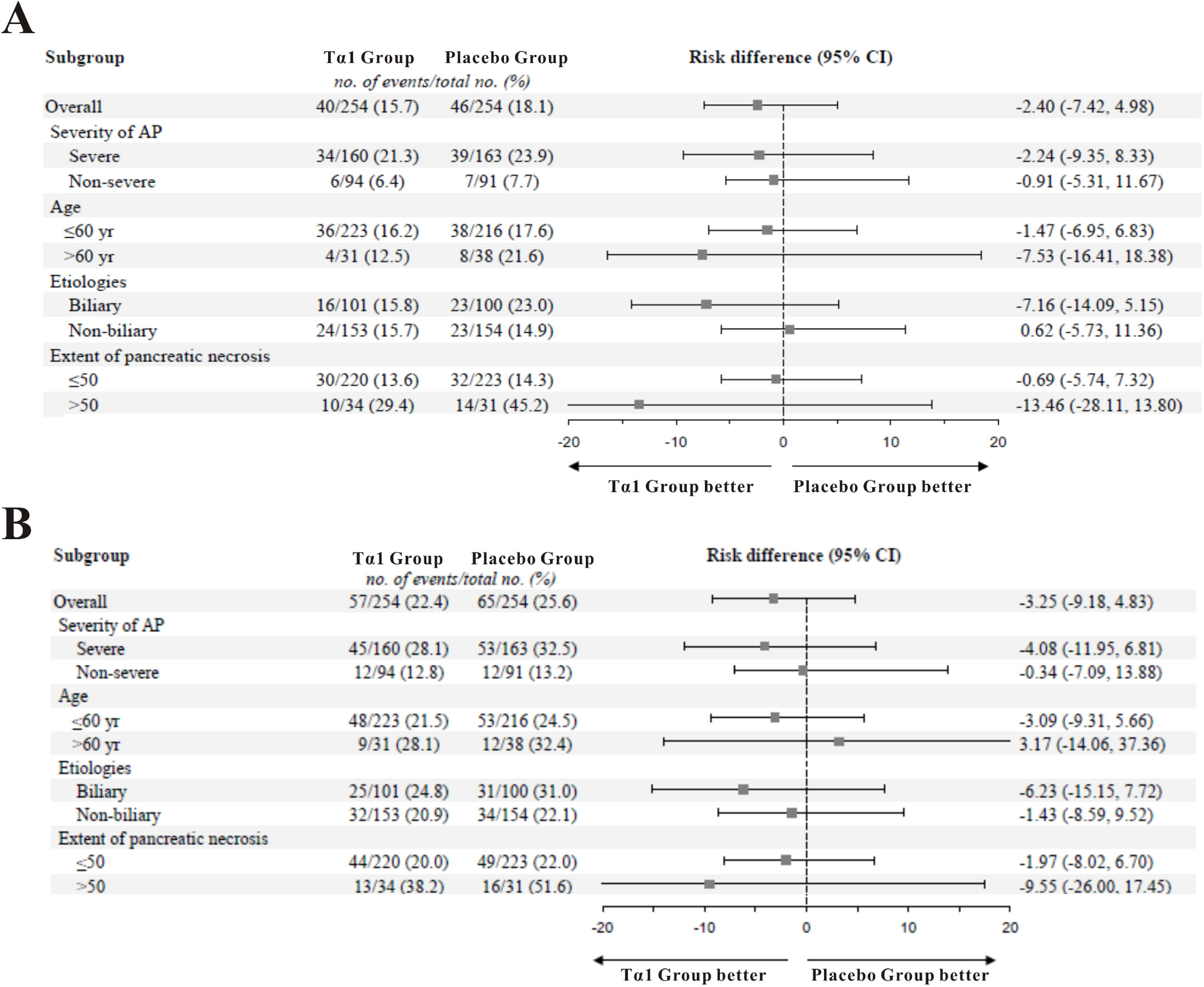
Subgroup analysis of the risk of infected pancreatic necrosis by the index hospital discharge and day 90. Panel A shows the relative risk of infected pancreatic necrosis during the index admission between the two treatment groups. Panel B shows the relative risk of infected pancreatic necrosis up to 90 days after randomization. A relative risk of less than 1.0 indicates better results for the Tα1 group

### Adverse events

Adverse events occurred in 21 patients in the Tα1 group and 19 patients in the placebo group (8.3 % vs. 7.5 %, P=0.742). The most common adverse event was venous thrombosis which occurred in 6 patients (2.4%) in the Tα1 group vs. 5 (2.0%) in the placebo group.

## Discussion

In this multicenter, double-blind, placebo-controlled, randomized trial, immune enhancement using Tα1 did not significantly reduce the incidence of IPN during the index admission or within 90 days of randomization in patients with ANP. Given the range of severity of AP ^2^, this study was designed to select more severe patients based on the APACHE II score at enrollment ^23^. However, we failed to show a difference in the primary outcome.

Our results are not consistent with the results from an experimental animal study ^17^ and the pilot clinical study ^24^. There are several possible explanations. First, current animal models can not recapitulate all aspects of human AP, especially for a complication such as IPN, which often occurs several weeks after admission ^25, 26^. Second, the pilot study recruited only 24 patients from a single center, making its findings vulnerable to type I error. Third, the dose regimen in the present trial is different from the pilot one with a longer duration of drug administration (one week in the pilot versus two weeks in the present) and lower initial dose (6.4mg per day in the pilot versus 3.2 mg per day in the present). There were two time-course considerations in designing the dose regimen: (1) infection mostly occurs beyond the second week after disease onset ^3, 27^, and a two-week regimen should cover the period interval better when prevention is possible; (2) immunosuppression typically develops early in the first week and usually slowly recovers during the second week ^12^, which is the reason for prescribing half the dose during the second week of treatment. A similar step-wise dose reduction was used in a previous study testing Tα1 in sepsis ^28^, showing that Tα1 could reduce 28-day mortality. The changes for initial dose were made because of concerns about the safety of the original dose regimen.

There is evidence to support a shifting balance between the systemic pro-inflammatory response and the compensatory anti-inflammatory response over the early course of AP ^13, 29^. It was considered that the pro-inflammatory response occurs in the first few days to weeks and that the compensatory anti-inflammatory response occurs later. However, analyses in patients with sepsis and AP suggest that these responses can also run in parallel and that there is an association between early-onset immunosuppression and poor outcomes in AP ^30, 31^. Previous trials investigating immunomodulatory therapy to block the early pro-inflammatory response have not been convincing ^32^, and this includes drugs like lexipafant ^33, 34^ and octreotide ^35^. In patients with severe COVID-19, observational studies showed that Tα1 attenuated lung injury and decreased mortality ^36, 37^. Despite the theoretical benefits of immune-enhancement with Tα1 and the encouraging results from the pilot study ^17^, Tα1 did not reduce the incidence of IPN or improve any of the clinical outcomes in this trial.

In the subgroup analyses, larger treatment effects were seen in patients with a greater extent of pancreatic necrosis (>50%) and those aged more than 60 years old, although not statistically significant. We should interpret all the subgroup results with caution. First, the power was not enough to detect the differences among treatments. Second, the definition of necrosis is relatively subject based on a single CT scan. Third, we excluded patients aged more than 70 years old given that age > or =70 has been proved to impact the clinical outcomes^38^, which makes the study subgroup for elderly patients even smaller.

In line with the excellent safety profile reported in previous studies, Tα1 showed satisfactory safety performance in this trial. Three patients discontinued treatment due to adverse reactions (one erythra and two unexplainable fever) but received the placebo.

The study has several limitations. The first is that the incidence of IPN may have been affected by the use of antibiotics, especially therapeutic, because this was not standardized but left to the clinical team to decide. The second is that there were problems (failed lab quality control) with the measurement of the monocyte human leukocyte antigen-DR (for monitoring drug effects) in half of the study subjects, and this could explain the negative result for this parameter. The third is that APACHE II misclassifies the severity of AP in almost a third of patients, which could also have contributed to the negative result ^39^. And lastly, the timing of treatment might have been too late. The current trial included patients up to one week after the advent of symptoms, which may increase the heterogeneity of the study population. Enrollment of a greater range of patients earlier in the disease course may have provided a better estimation of the agent’s effects. In addition to the timing of treatment, the appropriate duration of therapy is unclear.

In conclusion, the immune-enhancing Tα1 treatment of patients with predicted severe ANP (APACHE II ≥8 at enrolment) did not significantly reduce the incidence of IPN during the index admission compared with placebo. Future trials seeking to investigate this approach will need to determine the best way to select patients and decide on the most effective dose and duration of Tα1 treatment.

## Supporting information

Ethical Approval Document

## Data Availability

The data that support the findings of this study are available from the corresponding author, upon reasonable request.

## Abbreviations

ANP: acute necrotizing pancreatitis
AP: acute pancreatitis
CRP: C-reactive protein
GLM: generalized linear model
IPN: infected pancreatic necrosis
ITT: intention-to-treat
mHLA-DR: monocyte human leukocyte antigen-DR
PP: per-protocol
RAC: Revised Atlanta Classification
Tα1: Thymosin alpha 1

## Acknowledgment

This study was funded by SBE2016750187 of Science and technology project, Jiangsu Province, China, partly by SciClone Pharmaceuticals Holding Limited. The funders were not involved in the trial’s design, data collection, interpretation, or manuscript preparation.

We acknowledge the contribution of Mengjie Lu, Gang Li, Bo Ye, Yan Chen, Zhenping Chen, Youdong Wan, Miao Chen, Qingbo Zeng, Wei Zhao, Lening Ren, Dahuan Li, Qingcheng Xu, Keke Xin, Bing Xue, Hongguo Yang, Dongsheng Zhao, Feng Zhou, Zigui Zhu in the development and execution of this study.

