## Supplementary material for "Immune enhancement to prevent infected pancreatic necrosis: A double-blind randomized controlled trial": Ethical Approval Document

### Ethical Approval Document of TRACE trial

ID: 2015NZKY-004-02

**Project Name**: Thymosin alpha 1 in the prevention of infected pancreatic necrosis following acute necrotising pancreatitis (TRACE trial): protocol of a multicentre, randomised, double-blind, placebo-controlled, parallel-group trial

**Central center**: Jinling hospital

**Participating Centers**:

The First Affiliated Hospital of Nanchang University

The Affiliated Hospital of Qingdao University Medical School

The Affiliated Hospital of Zunyi Medical University

The Affiliated Nanhua Hospital, University of South China

The Affiliated Hospital 2 of Nantong University

The First Affiliated Hospital of Wannan Medical College

Shangqiu First People's Hospital

The 94th Hospital of PLA

Jiangsu Provincial Hospital of Integrated Chinese and Western Medicine

Zhejiang Provincial People's Hospital

Luoyang Central Hospital

The First Affiliated Hospital and College of Clinical Medicine of Henan University of Science and Technology

Clinical Medical College of Yangzhou University

Qilu Hospital of Shandong University

The First Affiliated Hospital of Anhui Medical University For the

Chinese Acute Pancreatitis Clinical Trials Group (CAPCTG)

**Major investigators:** Weiqin Li, Yin Zhu, Xinting Pan, Hong Mei, Chengjian He, Weili Gu, Weihua Lu, Shumin Tu, Haibin Ni, Guoxiu Zhang, Xiangyang Zhao, Junli Sun, Weiwei Chen, Jingchun Song, Min Shao, Jianfeng Tu

**Project classification**: Clinical Trials

**Ethical review Method**: By Meeting

**Review Date:** 2016-12-03

**Review comment:**

After review by the ethics committee, we agree to carry out this trial according to the revised study protocol and informed consent. Please carry out the clinical trials according to the original GCP and the protocol approved by the ethics committee to protect the rights and interests of involved patients. The ethics committee will follow up the trial until the end of the study.

The specific requirements are as follows:

(1) Whether major investigator changes or any modification of the clinical trial protocol /informed consent should be submitted for revision review application;

(2)Any serious adverse events must be submitted in a timely manner serious adverse events report;

(3) According to the specified annual/regular tracking review frequency, the research progress report must be submitted every year;

(4) In the process of the trial, if there is a violation of the protocol, violation scheme report should be submitted timely;

(5) If the applicant wants to suspend or terminate the clinical trial, the suspension/termination study report should be submitted in time;

6. Upon completion of the test, a final report should be submitted.

**Statement:**

The composition and working procedures of the ethics committee are in accordance with the requirements of the "drug clinical trial quality management standard (August 6, 2003)" issued by the China Food and Drug Administration and the guidelines for ICH clinical trials.

**The ethics committee of Jinling hospital**

**Contact person: Yuxiu Liu**

**Contact number :+86-025 80863**
